# Interstitial Lung Disease Fatigue & Breathlessness (ILD-FAB) Programme: A multidisciplinary feasibility study

**DOI:** 10.1101/2025.06.10.25329329

**Authors:** Jessica Mandizha, Rebecca Davies, Charlotte Crook, Anna Duckworth, Michael Gibbons, Joseph Lanario, Sarah Lines, Jessica Moss, Kate Taylor, Anne-Marie Russell

## Abstract

**Background:** Fatigue, breathlessness and cough are prevalent symptoms of Interstitial Lung Disease (ILD) adversely impacting quality-of-life and contributing to psychological distress. The Fatigue and Breathlessness (FAB) programme facilitates supported self-management for people living with life-limiting conditions such as cancer. We explore its utility when adapted for people living with ILD.

**Methods:** The 4-week ILD-FAB programme offers each group of up to 6 participants weekly two-hour sessions led by an ILD-specialist Physiotherapist and Clinical Nurse Specialist (CNS). Primary focus is on strategies to manage breathlessness, fatigue and wellbeing. Further, a 1:1 session with the ILD-CNS enables participants to set personalised goals and explore individual health beliefs/behaviours using a Cognitive Behavioural Therapy (CBT) assessment framework. The self-reporting Chronic Respiratory Questionnaire (CRQ-SR) evaluates breathlessness, fatigue, emotional function and mastery at baseline and after 4 weeks.

We facilitated eleven groups between March 2023 and December 2024.

**Results:** Forty-nine participants (26 male; median age 76 years [IQR=14]) were diagnosed with Idiopathic Pulmonary Fibrosis/IPF (n=21), Progressive Pulmonary Fibrosis/PPF (n=17) or non-progressive ILD (n=11) of various aetiologies. Lung function indicated a range of disease severity (FVC % predicted median: 70% [IQR=34])

Thirty-seven (76%) participants attended all four sessions, 6 (12%) attended three sessions, 2 (4%) attended two sessions and 4 (8%) attended one session.

Thirty-seven patients, all who attended at least 3 sessions, completed the CRQ-SR at baseline and Week 4. Fifty-nine percent of respondents (n=22) demonstrated clinically significant improvements in dyspnoea scores, 51% (n=19) in emotional functioning scores and 49% (n=18) in fatigue and mastery scores.

Thirty-five respondents (95%) demonstrated a clinically significant improvement in at least one domain.

All participants (100%) would recommend this programme to others.

**Conclusion:** These data demonstrate feasibility, acceptability and clinical effectiveness of an ILD-specific FAB programme. Further research will explore a range of outcome measures longitudinally in a larger cohort.

**Key Messages:**

- ***What is already known on this topic*** *-*The FAB programme is delivered in hospices and NHS trusts UK-wide to improve confidence in fatigue and breathlessness management and reduce anxiety for people living with life-limiting illnesses such as cancer. Formal evaluations are positive but limited by small sample sizes and the use of non-validated outcome measures.
- ***What this study adds*** *-*Our FAB programme, adapted for people with ILD, is feasible, acceptable and clinically effective.
- ***How this study might affect research, practice or policy*** *–* The FAB programme offers one multimodal approach to improving self-management in people with ILD. Further research in a larger, more heterogeneous patient population will optimise outcome measures, broaden acceptability and determine cost-effectiveness.

## Introduction

Interstitial Lung Diseases (ILDs) are a diverse group of serious respiratory conditions characterised by inflammation and/or irreversible fibrosis of lung tissue, causing frequently progressive breathlessness, fatigue and cough^1^. High symptom burden and psychological distress significantly and adversely impacts quality-of-life (QoL)^2^. Optimal management slows progression with anti-fibrotic medication and palliates symptoms to at least maintain, if not improve, QoL.

Supported self-management is an approach to improve people’s knowledge and ability to manage their own health^3^. It is a cornerstone of the UK NHS Long Term Plan’s commitment to make personalised care the norm, to increase autonomy, wellbeing and concordance and reduce unscheduled healthcare contacts^4^. Programmes are typically multimodal and include elements of education, peer support, symptom control and health coaching. ILD-related symptom burden and its impact on QoL is well documented^5,6^, as is the need for multidisciplinary approaches to improve symptom management, tailored education and effective psychosocial support^7,8^. Informational and supportive care needs, alongside patients’ desire to learn more about self-management approaches are defined as unmet needs^9-11^. The provision of appropriate interventions to reduce these unmet needs is a key health care priority^12^.

The mainstay of non-pharmacological interventions in ILD is Pulmonary Rehabilitation, a group-based education and exercise programme originally developed for patients with COPD and recommended in the British Thoracic Society (BTS) and National Institute of Health and Care Excellence (NICE) guidelines^13^. Uptake and completion of the programme is often low^14^. Other interventions to improve wellbeing in ILD include mindfulness^15-17^, where sadly, to date, low-quality science obscures our understanding of its effectiveness^18^. Singing^19-21^, Tai Chi^22^ and dancing^23^ offer alternative approaches, however the generalisability of these studies is difficult to determine due to heterogeneity of study design and small sample size.

The Fatigue and Breathlessness (FAB) programme is a complex, multimodal intervention delivered in hospices and acute NHS trusts for people living with conditions such as cancer and heart failure. FAB programmes have organically evolved and are currently delivered in a non-standardised way but do include education and group discussion on approaches to managing breathlessness, panic and fatigue. The aim of FAB is consistent: to improve participants’ ability to self-manage their symptoms.

Despite its widespread adoption and implementation, the evidence base for the FAB programme is limited and the lack of standardisation is problematic for comparative evaluation. Our scoping review of the literature did not identify any full reporting of original research on the FAB intervention. Six published conference abstracts were identified reporting small-scale evaluations of FAB between 2016 and 2023^24-29^. Outcome measures included the Chronic Respiratory Disease Questionnaire (CRQ) and Integrated Palliative Care Outcome Scale (IPOS), reporting some positive changes. Notably details of the content and delivery of the programme, participant numbers and their demographics were often incomplete. One abstract reporting CRQ data on ten participants noted deterioration in certain domains (1 patient in Dyspnoea domain, 1 patient in Emotional Functioning domain and 5 patients in the Mastery domain). This may suggest programme limitations, unintended consequences, seasonal factors, disease progression or unsuitability of the outcome measure^28^.

Multimodal interventions with a focus on symptom control, quality of life, peer support and psychosocial input are likely to have a positive impact upon patients with respect to their supportive care needs^12^. To explore the utility of delivering a FAB programme adapted for people living with ILD, we conducted a feasibility study.

## Methods

Having identified patient need and the gaps in the literature, we convened a multidisciplinary clinical academic group to discuss the development and implementation of an ILD-FAB programme. We sought perspectives from patient-partners. To enhance the original FAB programme for an ILD population, priority topics identified from within the group and relevant literature included pathophysiology of ILD, managing weight loss and optimising the use of oxygen therapy. From this mind mapping exercise, an initial framework of core content was agreed. To allow for future development, the participant feedback survey specifically invited participants to identify any additional topics they would like covered.

### Population

The ILD- FAB programme was offered to all patients over the age of 18 years who were able to consent, during routine clinic visits or phone calls at a regional NHSE specialist commissioned ILD service in the UK. As this feasibility study was delivered face-to-face, participants needed to live within the hospital catchment area, be able to travel and intend to attend the full course. A follow up phone call was made to those expressing an interest to discuss the programme in more detail, identify any specific needs and confirm dates and times (RD).

### Intervention

The FAB-ILD programme ran in a community hospital over 4 consecutive weeks with up to 6 participants per group. Importantly for this group of people with breathlessness, on-site car parking was available. Each two-hour session was led by an ILD-specialist Physiotherapist (RD) and ILD Clinical Nurse Specialist (CNS) (JM). Ground rules establishing parameters for confidentiality and mutual respect were agreed at the first session to promote trust and an emotionally safe environment.

The sessions began with a taught element focussing on strategies to self-manage symptoms (Table One), including a session on optimising dietary intake, delivered by an ILD-specialist dietitian (KT).

**Table One:**
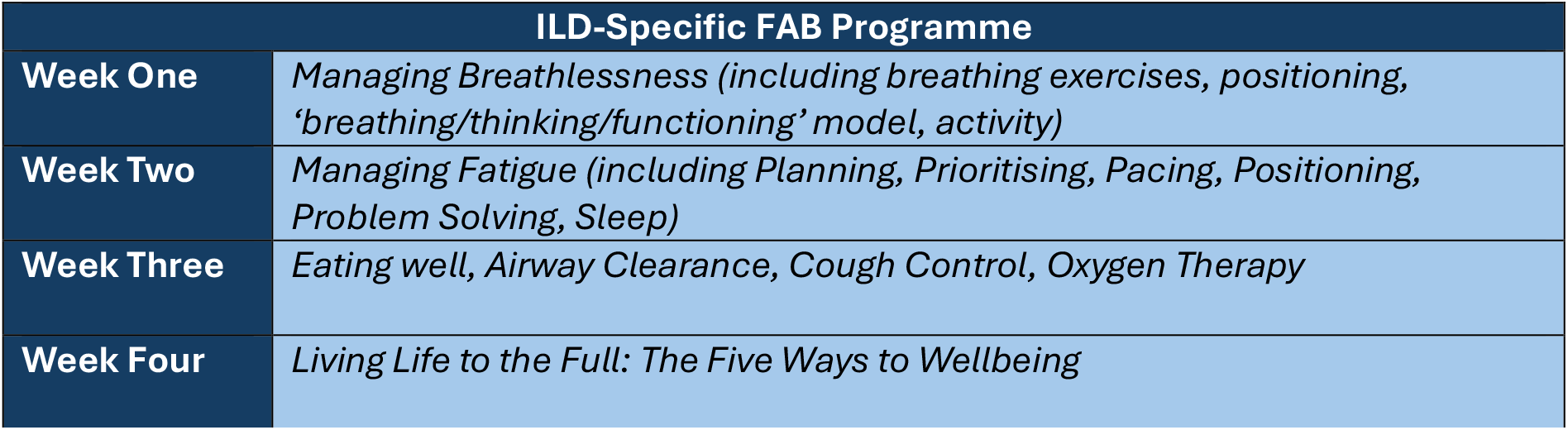
Topics taught within FAB programme.

Group discussion followed, with refreshments, offering participants an opportunity to consolidate their learning, ask questions and offer each other peer support. This was a key aspect of the programme.

The second half of the session supported participants in practical exercises to improve understanding and increase confidence. This group work included energy mapping, relaxation techniques and chair-based exercise.

Over the four-week programme, participants received a one-to-one session with the CNS to explore individual health beliefs, behaviours or issues and, if relevant, set personalised goals. These conversations were captured using the Cognitive Behavioural Therapy (CBT) ‘hot-cross- bun’ model, which diagrammatically illustrates how thoughts, emotions, physical feelings and behaviours interact with each other within a given situation^30^ (Figure One). This tool was selected as it is also reflective of the ‘Breathing, Thinking, Functioning’ model which is introduced to participants within the programme to help explain the multidimensional nature of breathlessness^31^.

**Figure One:**
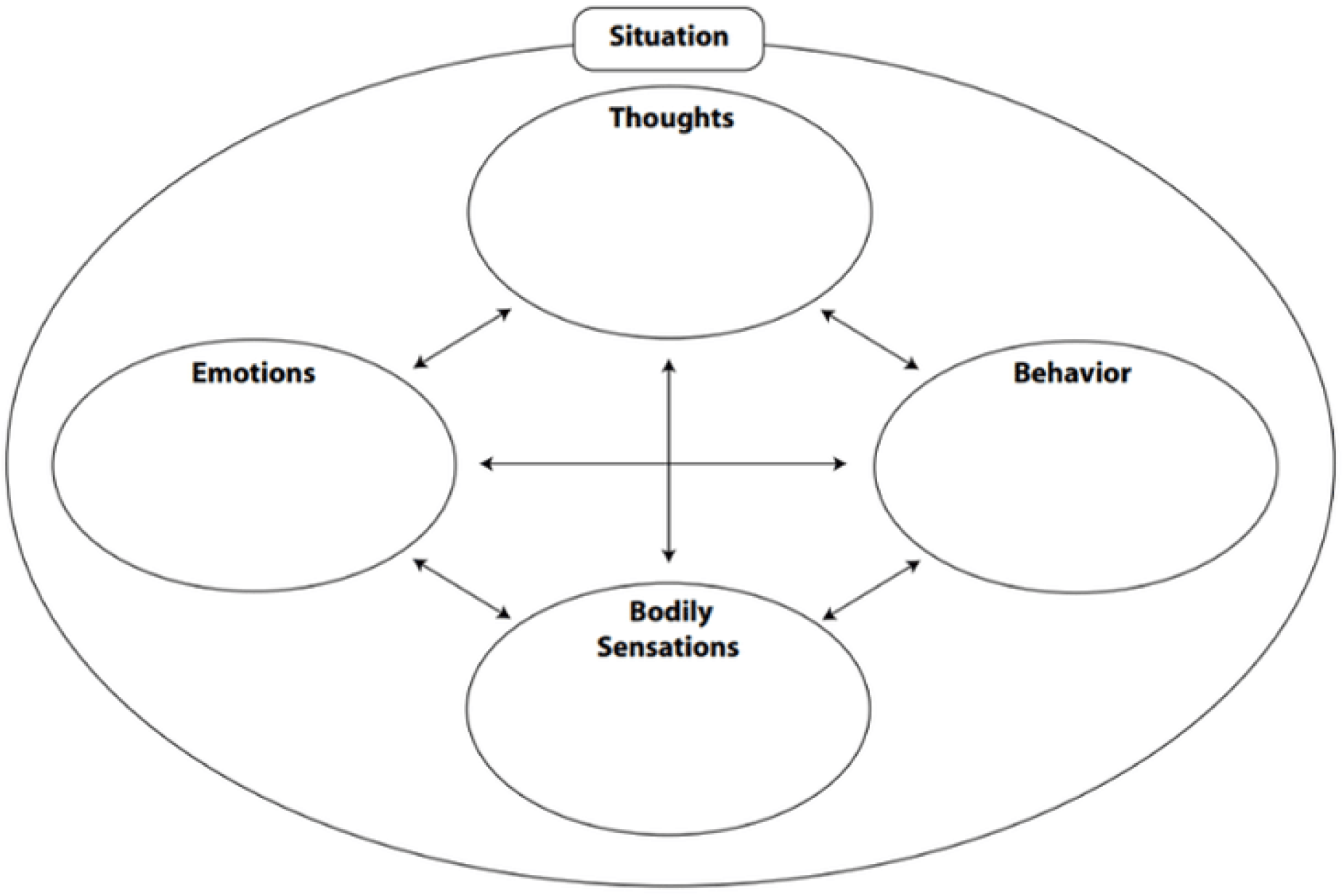
CBT ‘Hot-Cross-Bun’ Model.

Participants received a follow up phone call from one of the programme leads (JM or RD) approximately 6 weeks after completion. The aim of the call was to reiterate the strategies discussed within the programme, re-evaluate goals, identify any outstanding needs and ensure relevant referrals, for example, to Pulmonary Rehabilitation or hospice services, were in place. Any participants who had not managed to complete the programme were also followed up at this point.

### Outcome Measures

Participants were invited to complete a self-reported Chronic Respiratory Questionnaire (CRQ- SR) on arrival and at the end of Week 4. The CRQ is designed to measure health-related quality of life (HRQOL) in people with chronic respiratory disease. It evaluates four aspects: dyspnoea, fatigue, emotional function and mastery. Each domain includes 4 to 7 items, with each item graded on a 7-point Likert scale. Item scores within a domain are summated to provide a total score for each domain. Higher scores indicate better HRQOL. The 4 domains are scored separately, illustrating changes in individual domains of HRQOL^32^.

A minimal clinically important difference (MCID) of 0.5 per item is widely accepted for the CRQ- SR^32-35^. However, it is important to consider that patient-determined MCIDs are typically associated with smaller change to scores than physician/expert determined MCIDs, meaning the patient benefit of an intervention may be underestimated.

The CRQ-SR has demonstrated good test-retest reliability and has been shown to be more sensitive than other measures in detecting small changes in HRQOL over relatively short periods of time^36^.

### Survey

On completion, participants were invited to complete a semi-structured survey on their experiences, inviting comments on relevance and delivery of content and length of course. They were also asked to define one thing from the course that made a difference to their symptom management. The survey was posted to participants who could not or did not attend the final session with a return envelope, to try and capture a broad range of opinions and experiences.

Returned survey forms were read repeatedly for familiarity, with emergent patterns identified inductively and themes generated (JM). Data were scrutinised, compared, discussed and ratified by AMR and RD.

### Ethical Considerations

The project was registered, peer-reviewed and approved as a service evaluation project [Ref: 24- 1599] by an NHS Respiratory Speciality Governance Group at the host Trust.

## Results

### Participants

Demographic data of 49 participants is detailed in Table Two.

**Table Two:** Participant demographics.

| Demographics |  | Number |
| --- | --- | --- |
| Gender | Male | 26 |
|  | Female | 23 |
| Age | <50 years | 4 |
|  | 50-59 years | 0 |
|  | 60-69 years | 9 |
|  | 70-79 years | 22 |
|  | 80-89 years | 14 |
| Diagnosis | Idiopathic Pulmonary Fibrosis (IPF) | 21 |
|  | Progressive Pulmonary Fibrosis (PPF) | 17 |
|  | Non-progressive ILD | 11 |
| Baseline FVC% predicted | >80% | 19 |
|  | 70-79% | 6 |
|  | 60-69% | 10 |
|  | 50-59% | 8 |
|  | 35-49% | 6 |
|  | <35% | 0 |

### Attendance

Thirty-seven (76%) participants attended all four sessions, 6 (12%) attended three sessions, 2 (4%) attended two sessions and 4 (8%) attended one session. Absence was frequently reported to be due to acute illness or concurrent health appointments.

### Outcome Measures

Thirty-seven participants (76%) completed the CRQ-SR at both timepoints. Eight participants (16%) who did not attend the last session completed the initial CRQ-SR. Four participants (8%) completed the questionnaire incorrectly, so results were invalid.

Thirty-five respondents (95%) demonstrated a clinically significant improvement in at least one domain. Twenty-two respondents (59%) demonstrated clinically significant improvements in dyspnoea domain scores, 19 (51%) in emotional functioning domain scores, 18 (49%) in fatigue domain scores and 18 (49%) in mastery domain scores (Figure Two).

**Figure Two:**
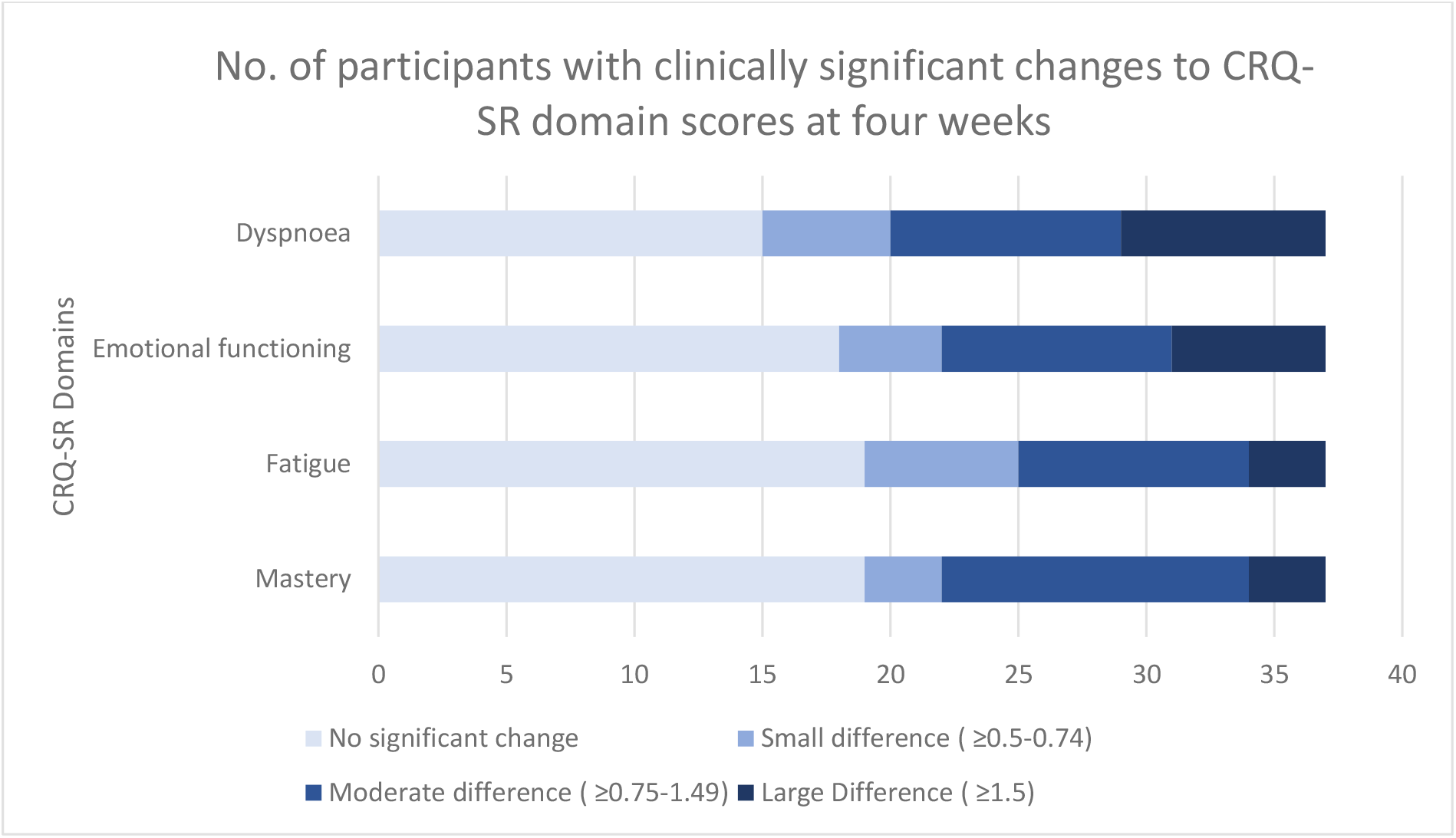
No. of participants with clinically significant changes to CRQ-SR domain scores at four weeks.

### Participant Feedback

Forty-eight participants completed a feedback survey. Forty-five participants (94%) felt the 4- week course seemed the right length. Two participants (4%) felt it should be longer, and 1 participant (2%) felt it should be shorter.

100% of participants felt the subject matter was relevant and delivered in a way that was easy to understand. 100% of participants would recommend this course to other people who experience breathlessness.

When asked to note any relevant topics not covered within the programme, eleven participants commented, adding topics such as normal lung function, medication management, managing deterioration and end-of life planning.

Analysis of the qualitative data identified three main themes: 1) Increased optimism; 2) Significance of peer support; 3) Impact of facilitator characteristics. The themes are discussed briefly here, alongside illustrative quotes (Table Three).

**Table Three:**
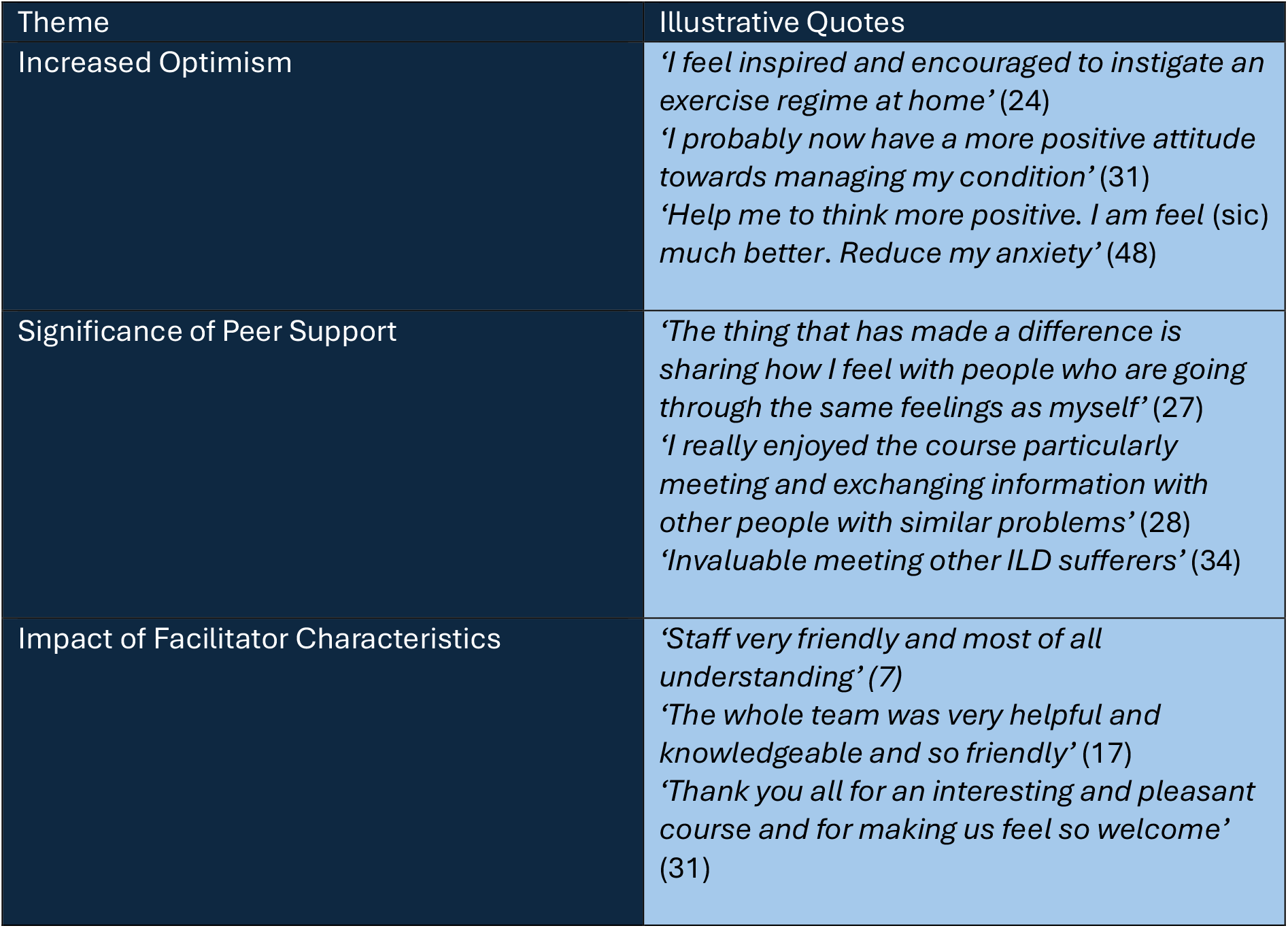
Themes with illustrative quotes (Participant number)

### Theme One: Increased optimism

Beyond gaining practical tools, participants expressed increased positivity, confidence and motivation following the course.

### Theme Two: Significance of peer support

Meeting and sharing with others with similar lived experiences was highly valued. For some participants, this was the first time they had met others with the same diagnosis.

### Theme Three: Impact of facilitator characteristics

Specialist knowledge and understanding was important to participants, alongside a friendly, welcoming approach.

## Discussion

Thirty-five patients (95%) demonstrated a clinically significant improvement in at least one domain of the CRQ-SR, particularly in the dyspnoea domain. The ILD-FAB programme was positively evaluated by participants. Retention was high, with 37 patients (76%) attending all four sessions.

A UK-based supported self-management programme for people with ILD has not previously been reported on, despite representing an unmet need. Our pilot data demonstrates initial acceptability of an ILD-FAB programme and suggests that such a programme may effectively improve health-related quality of life for participants in the short-term. Providing detailed information about how this is delivered may allow others to adopt the programme. Equally, it may need to be adapted to suit other teams or meet particular needs of local patient groups.

Themes arising from the feedback survey suggest that it may be important to replicate peer support if a different delivery method is chosen beyond face-to-face, such as a self-directed or digital route. Facilitator characteristics, such as specialist knowledge and good interpersonal skills, may be as important to the success of the programme as professional background.

The FAB programme represents an additional tool to support self-management approaches in ILD and has the potential to be used as an alternative or precursor to Pulmonary Rehabilitation. In other therapeutic fields, increased patient choice has demonstrated a positive impact on retention and adherence^37^.

### Limitations

This was a feasibility study and so the data represents a relatively small number of participants. Only one patient-reported outcome measure (PROM) was used and there is some missing data. The CRQ-SR was selected as it captures the domains which the FAB programme specifically seeks to address, and includes mastery, which is key to self-management approaches. The CRQ has been reported on in a previous evaluation of FAB^28^ and is commonly used to measure the impact of Pulmonary Rehabilitation programmes, enabling comparison. Whilst the CRQ-SR has the sensitivity to capture small changes, even over a short period of time, its slightly complex presentation increases the risk of incomplete data.

Alternative PROMs may be more appropriate and warrant further exploration, with input from patient-partners to minimise the burden. These may include disease-specific measures, such as the IPF-PROM, which is a12-item IPF-specific measure of health status, measuring physical and psychological experiences of breathlessness, emotional well-being and energy levels^38^. Symptom-specific measures may also be useful, such as the Dyspnoea-12, which is a 12-item questionnaire that assesses breathlessness and has been previously used and validated in ILD^39- 40^. In addition, functional measures such as daily step count may represent a meaningful outcome to patients.

As a feasibility study to explore proof of concept, a single centre was selected. This excluded outlying patients, meaning more geographically or socially isolated patients may not have been able to access the programme. This will need to be considered carefully in subsequent work so as not to potentially widen inequities in health care provision. The cohort for this centre is predominantly white. Results may therefore not be reflective of a wider, more ethnically diverse ILD population. Patient-reported outcome measures were completed immediately following the programme at the end of the Week 4 session. Longer-term impact or any decreasing effect of the programme is therefore not captured within the scope of this project.

### Recommendations

Accessing face-to-face services is particularly challenging for frail, rural or underserved populations^41^, those with financial constraints or less English language fluency. Related health inequalities have been observed in ILD^42^. Further research and development of the FAB programme should include considerations for these patient groups to increase inclusivity. This may include considering alternative methods of delivery which do not involve travel, translation of programme material into other languages, conducting research in a more ethnically diverse geographical area and actively targeting global majority participants.

The role of informal carers in self-management is key and the impact of ILD on family members is well recognised^43^. Future work must account for this and capture the experience of informal carers and the impact of the programme on them, including both those who attend FAB with the patient-participant and those who do not.

In-depth, qualitative data will enrich our understanding of both participant and carer experience, contributing to the development of future programmes as well as informing decisions about ongoing research design.

Supported self-management of chronic illness is a long-term endeavour which is particularly complex in the context of a progressive illness. Longitudinal work in a larger diverse cohort, utilising a broad range of outcome measures will give greater understanding of the FAB programme for patients living with ILD.

## Conclusion

Thirty-five patients (95%) demonstrated a clinically significant improvement in at least one domain of the CRQ-SR. The FAB programme was positively evaluated, with all participants stating they would recommend to others. Retention was high, with 37 patients (76%) attending all four sessions. Further research is needed to establish effectiveness in a larger, more diverse patient population.

## Data Availability

All data produced in the present study are available upon reasonable request to the authors

